# Effects of the Variance in Prevalence of the DASH Diet in Nursing Homes on Seniors With Elevated Blood Pressure or Hypertension

**DOI:** 10.1101/2022.09.18.22280072

**Authors:** Ryan Liu, Tsz-Kiu Chui

## Abstract

The DASH diet is a dietary pattern designed to help treat or prevent hypertension. The diet includes foods rich in potassium, calcium, and magnesium while limiting foods high in sodium, saturated fat, and added sugars. The DASH diet’s relationship with hypertension in adults is well defined, but the effect of its variance in prevalence in nursing homes on seniors is not. We performed a mixed-methods observational study incorporating a survey collecting anonymized nursing home data regarding the number of servings of various food groups provided to senior residents per day. The data were analyzed using the Fung et al. DASH diet scoring method. We then conducted an interview with nursing home dieticians to investigate the public health effect of the DASH diet on hypertension in residents. Lastly, a content analysis was performed of nursing home menus to support the data from the surveys. In a pool of 11 nursing homes, 100% of the facilities incorporated some aspect of the DASH diet, and in a pool of five nursing homes, 60% achieved high adherence to the DASH diet. We additionally confirmed a previously observed negative relationship between red meat consumption in seniors to higher risk and poorer prognosis of hypertension. These analyses bolster the DASH diet’s potential role as a hypertension prevention mechanism in nursing homes, suggesting that future DASH diet interventions may hold promise.

## 2. INTRODUCTION

This paper explores the prevalence of Dietary Approaches to Stop Hypertension (DASH) diet elements in Central Texas nursing homes that care for residents with hypertension and how each respective adherence level contributes to the blood pressure and holistic health of nursing home residents.

High blood pressure is a common problem in the United States with 24% of the US population having hypertension and less than half of the population having optimal blood pressure [2]. A systolic pressure below 120 mmHg and a diastolic pressure below 80 mmHg is considered normal blood pressure for an adult [18].

It has been almost 25 years since the DASH diet study’s results were published in July of 1997 and an important milestone was crossed in the study of nutritional science. Sponsored by the US National Institutes of Health, the DASH diet limits saturated and trans fat, while increasing the intake of potassium, magnesium, calcium, protein, and fiber. These nutrients have been shown to help control blood pressure [10]. Although the diet has proved highly effective in lowering blood pressure in hypertensive patients, national adherence to the diet is low. Less than 1% of the US population was fully adherent to DASH in 1988 to 2004, and this trend continues today [17]. With hypertension being a main risk factor for heart disease and stroke, spreading the DASH diet is critical to improving public health in the US.

## 3. LITERATURE REVIEW

### Reduced Blood Pressure Via the DASH Diet

The two studies written about are the initial proposed DASH diet study (study one) and the reduced-sodium diet combined with the DASH diet study (study two and study three). In both studies, researchers have concluded that there is a correlation between reduced blood pressure and adherence to the DASH diet.

The original DASH diet study included adult subjects, two-thirds of which were racial or ethnic minorities, with SBP of less than 160 mmHg and a diastolic blood pressure of between 80 to 95 mmHg [2]. In the second study, the researchers followed up on the original DASH diet trial and compared the effects of low versus high sodium, DASH versus control, and both (low sodium-DASH vs. high sodium-control diets) on systolic blood pressure [11]. Study two builds on the original study by adding the variable of sodium intake.

Study one was conducted in three phases: screening, run-in, and intervention. The control diet tested included elements present in a typical American diet, while the combination diet was high in fruits, vegetables, and low-fat dairy foods and had low amounts of saturated fat, total fat, and cholesterol [2]. The trial’s primary outcome was change in diastolic blood pressure, while the secondary outcome was change in SBP [2]. The participants of study two had the same qualifications and the trial had the same primary outcomes and the same measurement tactics as those of the original DASH diet study. The participants of study two were divided into DASH or control diets and fed either low, medium, or high sodium levels and were fed that diet for 30 days. The high level reflected the average salt intake of Americans, the medium level reflected recommended intake, and the low level reflected a level below the recommended intake [11]. This is building on study one’s methods by including the factor of sodium intake.

The results of study one found that the combination diet reduced SBP by 5.5 mm Hg more and diastolic blood pressure by 3.0 mm Hg more than the control diet did [2]. In study two, the results found that reducing sodium intake from high to low levels for the participants assigned to the control group resulted in SBP reductions shown in the chart below [11].

Reducing sodium intake from high to low levels for the participants assigned to the DASH diet resulted in SBP reductions shown in the chart below [11].

### Impact of Variance in Sodium Intake on Blood Pressure

Study three’s participants had the same qualifications as those of study one and two; study three also had the same primary outcome and used the same methods as those of study 2. The third study concurred with study two, finding that the combined effects the DASH diet and a low sodium intake were substantially greater than the effects on blood pressure than either intervention alone [15].

In all three studies, the reductions in blood pressure in subjects with hypertension were more significant than those in subjects without hypertension and were comparable to those observed in trials of drugs for hypertension [2]. In study two, the combination of low sodium intake and the DASH diet reduced SBP, with greater reductions at higher levels of baseline SBP. At the high sodium level, the DASH diet was compared with the control diet, and the former lowered SBP in each stratum. At the low sodium level, the effects of the DASH diet in the four strata of baseline SBP were not significant [11]. Since there is a 10 mm Hg difference in final SBP between hypertensive patients in the DASH diet with low sodium group and those in the control diet with high sodium, both studies show that hypertensive patients can lower SBP through dietary interventions alone.

### Elements of the DASH Diet Leading to Reduced Blood Pressure

The DASH diet emphasizes vegetables, fruits and low-fat dairy products, whole grains, fish, poultry and nuts and encourages a reduction in sodium intake [13]. Scientists have recently found a potential reason using metabolites: substances produced during digestion. The study was done on 64 African Americans with hypertension [1]. All of the participants were assigned to a reduced-sodium diet. The researchers found that reducing sodium intake increased two levels of metabolites: both correlated with low blood pressure levels and arterial stiffness [1]. These two metabolites are called beta-hydroxyisovalerate and methionine sulfone. Although further studies are needed to confirm the correlation, this is one of the first studies to investigate the reason behind reduced sodium intake and lowered blood pressure.

### Adherence Strategies for the DASH Diet

Although long-term adherence to the DASH diet has been proven to lower blood pressure levels in hypertensive individuals, those who claim to fully adhere to the DASH diet only adhere 60% to the recommended food intake levels [7]. This is an issue because participants are not receiving the full benefit of the diet. Their magnitude of blood pressure lowering would be markedly less than someone of the same qualifications who adheres 100% to the DASH diet. Level of adherence to a diet is highly influenced by how similar the “new” diet is to the individual’s usual diet [9]. Making the diet flexible and adaptable to the person’s usual diet, which can be done with the DASH diet, as it only controls food-type intake, can increase the overall adherence to the DASH diet [9].

### Gap in the Research

Research about the DASH diet up to this point has focused on testing the efficacy of the diet [2]. As more research suggests that the DASH diet is indeed effective in lowering blood pressure, especially in those with high-strata hypertension, more research is needed about the prevalence of the DASH diet [2].

The current gap in the research lies in researching more about how often nursing homes that care for hypertensive patients incorporate the DASH diet into their menus. It is critical that facilities that care for hypertensive patients are cognizant of the beneficial effects of the DASH diet on the health of their residents. Since the complete adherence rate to the diet is so low, being aware of specific foods that are “DASH diet friendly” is important. Even if nursing homes do not fully adhere to the DASH diet, incorporating certain elements of the DASH diet, such as more leafy vegetables, can have benefits: although less than those when fully adhering to the DASH diet, they can nevertheless have a positive impact on the health of their residents. In order to address this gap in the research, the guiding research question is: How often are elements of the DASH diet incorporated into the menus of Central Texas nursing homes that care for hypertensive patients?

## 4. RESEARCH DESIGN AND METHODOLOGY

### Study Design

This study seeks to analyze the prevalence of elements of the DASH diet in Central Texas nursing homes that care for residents with hypertension. Its goal is not only to make nursing homes aware of how effective their menus are in helping their patients control their blood pressure, but also to show them how they can improve their menus to fill any nutritional gaps they may have. Increasing their familiarity with the DASH diet will help them recognize that high blood pressure can be counteracted with DASH diet foods.

The study was conducted in three parts. Both quantitative and qualitative methods were used to analyze DASH diet adherence. First, a survey was conducted asking numerical questions about how many servings of different food groups they provide to their residents per day. These questions were based on the Fung et al. DASH diet scoring method. This was a quantitative analysis method and helped to gather preliminary data about participants’ DASH diet scores. It also allowed for more insight into which specific diet they used, if any, and gave the opportunity to ask for their menus and request an interview. An interview was then conducted with nursing home dieticians asking about if they incorporate the DASH diet into their menus. This was a qualitative analysis method that helped foster a better understanding of how they design their facility’s menus. A content analysis was also conducted of nursing home menus. This study design is important because it combines both quantitative and qualitative data points that helped clarify how nursing homes incorporate elements of the DASH diet into their residents’ food. Not only can the numbers be understood, but also the reason and logic behind those numbers.

### Subjects

The subjects of this study included chefs, nutritionists, and dieticians. These are the individuals who are responsible for designing the diets that residents are served, monitoring the health of the residents, and adjusting the menu according to the health data that they gather. They are the most appropriate population to study, as they are certified in the field of nutrition and have the most oversight of their particular nursing home’s menu. They would also have the most precise numbers and most detailed reasoning because of this. Potential subjects of this study were gathered from a search of nursing homes in the Central Texas area. Each facility was individually contacted to make sure that they housed hypertensive residents.

### Measurement Indexes

The survey in this study was based on “Fung’s DASH diet index” from Fung et al. [8]. The utilization of this tool was granted permission by Fung et al. The scoring system is based on quintiles with the lowest intake of health components receiving one point and the top quintile of health components receiving five points. The score for unhealthy foods is reversed [16]. The score ranges from 8 (lowest adherence) to 40 (highest adherence). The scoring is based on average food intake with options from “never or less than once per month” to “six or more times per day” for each food item [8]. The foods emphasized in this scoring system are high intake of fruits, vegetables, nuts and legumes, low-fat dairy products, and whole grains and low intake of sodium, sweetened beverages, and red and processed meats [8]. To have high adherence to the DASH diet, all five points must be received for 60% of the food groups.

### Serving Size Requirements for Each Food Group Based on DASH Diet

Note that quintiles one and five of sodium, red and processed meats, and sweetened beverages are reversed so that the lowest quintile receives five points and the highest quintile receives one point. This scoring system gives a quantitative analysis of diets and can produce correlation coefficients, or a measure of the strength of the relationship between two variables, between diet and hypertension.

Although the original subjects of this index were women at risk of coronary heart disease and stroke and had never been used for seniors, the purpose of measuring adherence to the DASH diet was consistent in both cases. To accommodate the study, supplemental questions were added to the survey. These questions were all reviewed and approved by the Institutional Review Board (IRB). The questions collected qualitative data such as if they were aware of or provided meals based on the DASH diet or if they provided meals based on any diet. The menus that were provided to their residents as well as an interview with their nutritionist, dietician, or chef were requested.

### Procedures

After initially gathering a list of potential subjects and their facility’s phone number from doing an online search of nursing homes in the Central Texas area, each facility was contacted once. If they did not respond, they were left a voicemail and were contacted another time a week later. If they did respond and agreed to the study, the dietary manager would be asked the questionnaire questions over the phone. Five facilities responded to the survey and twelve provided menus.

The twelve menus that were provided by nursing home facilities were analyzed using a spreadsheet. Each row had a nursing home code name and each column had a food that fit in the sheet’s specific category. There were seven categories: fruit, vegetable, nuts and legumes, whole grains, low-fat dairy, red and processed meats, and sweetened beverages. Every time a nursing home’s menu served a food that was mentioned in the spreadsheet, the food was marked under the nursing home’s name. This helped organize the different foods that each nursing home served and see trends in the data.

The dietary managers that agreed to participate in a survey were asked a series of questions about their facility’s menu. Each facility was also asked one interview question, asking whether they intentionally incorporated the DASH diet in their facility’s menus. Each facility was kept anonymous in the survey to prevent privacy issues and each facility is referred to as their classified number. The study design and procedures were also both approved by the IRB.

### Delimitations

To gather a list of the most specific and suitable subjects, only nursing homes who housed hypertensive residents were contacted. Also, only nursing homes in the region of Central TX were contacted to narrow the study to a specific area that has a high proportion of hypertensive seniors.

## 5. RESULTS

### Quantitative Results

The questionnaire was answered by three facilities. Table 3 portrays the frequency of facilities that met the criteria based on the DASH diet scoring method by Fung et al. for each question/food group. To meet the criteria, facilities must score five points (highest adherence level) on the DASH diet scoring method. Along with each question is the criteria to score five points on the DASH diet scoring method. For example, to score five points on the DASH diet scoring method and to meet the criteria for question 1, the facility must serve at least four servings of fruit per day.

**Table 1.** Blood pressure changes measured in mm Hg.

| Reducing sodium intake from high to low levels for participants assigned to the control diet | Reducing sodium intake from high to low levels for the participants assigned to the DASH diet |
| --- | --- |
| -3.20 mm Hg | -5.3 mm Hg |
| -8.56 mm Hg | -7.5 mm Hg |
| -8.99 mm Hg | -9.7 mm Hg |
| -7.04 mm Hg | -20.8 mm Hg |

**Table 2.** Lower end of the serving size represents quintile 1 of the DASH diet criteria and higher end of the serving size represents quintile 5 of the DASH diet criteria.

| Food Group | Quintile One-Five Per Day |
| --- | --- |
| Fruits | 0.7 servings to 4.1 servings |
| Vegetables | 1.1 servings to 4.6 servings |
| Nuts and legumes | 0.3 servings to 1.5 servings |
| Whole grains | 0.1 servings to 2.4 servings |
| Low-fat dairy | 0.1 servings to 2.3 servings |
| Sodium | 1041 mg to 2676 mg |
| Red and processed meats | 0.4 servings to 1.8 servings |
| Sweetened beverages | 0 servings to 1.2 servings |

**Table 3.** n=5.

| Question with Criteria to Number of Points on the DASH Diet Scoring Method by Fung et al. | Frequency of Facilities that Met Criteria (%) | Frequency of Facilities that Did Not Meet Criteria (%) |
| --- | --- | --- |
| How many fruit servings do you provide per day? (Criteria = at least 4 servings) | 4 (80%) | 1 (20%) |
| How many vegetable servings do you provide per day? (Criteria = at least 5 servings) | 5 (100%) | 0 (%) |
| How many servings of nuts and legumes do you provide per day? (Criteria = at least 2 servings) | 3 (60%) | 2 (40%) |
| How many servings of whole grains do you provide per day? (Criteria = at least 2 servings) | 3 (60%) | 2 (40%) |
| How many servings of low-fat dairy do you provide per day? (Criteria = at least 2 servings) | 2 (40%) | 3 (60%) |
| How many milligrams of sodium do you provide per day? (Criteria = less than 1041 mg) | 0 (0%) | 5 (100%) |
| How many servings of red and processed meats do you provide per day? (Criteria = less than 0.4 servings) | 1 (20%) | 4 (80%) |
| How many servings of sweetened beverages do you provide per day? (Criteria = 0 servings) | 3 (60%) | 2 (40%) |

### Frequency of Criteria Met/Not Met through Questionnaire

From the data in table 3, the total number of diet elements from facilities that met the criteria (21) was greater than the number that did not meet the criteria (19) for each question/food group. If each of these facilities were to complete the DASH diet scoring method, the majority of them would be awarded the highest adherence level score. The top two food groups that met the criteria and had the highest possible adherence score were vegetables (100%; Q2) and fruit (80%; Q1). The top two food groups that did not meet the criteria were sodium (100%; Q6) and red and processed meats (80%; Q7).

### Frequency and Percentage of Criteria Met/Not Met from Total Criteria by Facility

Table 4 depicts the frequency and percentage of criteria that were met and not met from total criteria, organized by facility. Facilities A, C, and E (60%) had a total score of at least five out of 8, making these facilities have a high adherence rate to the DASH diet. These facilities’ foods aligned most with the DASH diet. Facilities B and D (40%) had a total score of less than five out of 8, making them have a low adherence rate to the DASH diet. These facilities’ foods did not align with the DASH diet. Although none of the nursing homes completely adhered to the DASH diet, each facility had at least three food groups that did adhere to the DASH diet guidelines.

**Table 4.**
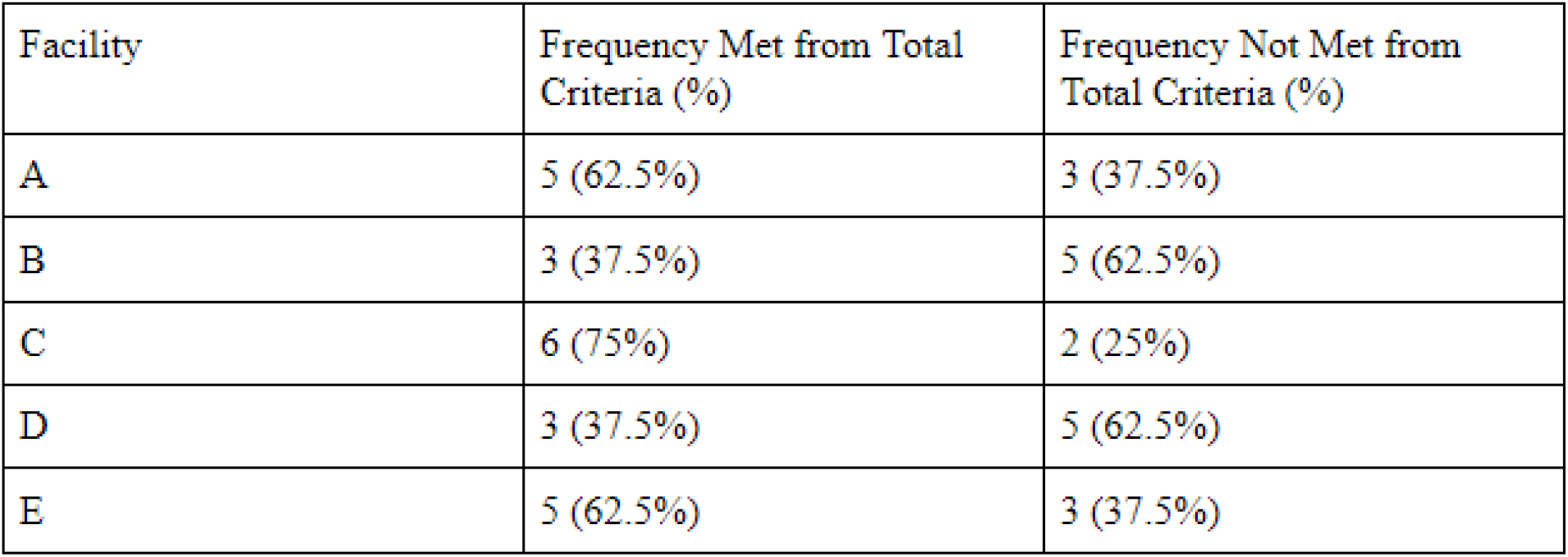
n=5. Based on raw data from Table 3.

### Frequency and Percentage of Adherence Levels for All Facilities

Table 5 consolidates the results from Table 4, showing the frequency and percentage of different adherence levels for all the facilities. The majority (60%) of facilities highly adhered to the DASH diet, while 40% of the facilities had low adherence to the DASH diet.

**Table 5.** Based on raw data from Table 4.

| Adherence Level | Frequency (%) |
| --- | --- |
| High Adherence | 3 (60%) |
| Low Adherence | 2 (40%) |

### Qualitative Results

#### Interview

During the questionnaire, one interview question was asked: do you intentionally incorporate elements of the DASH diet into the facility’s menu? All five of the facilities responded “no”. All five facilities also provided a reason for their answer, citing their ignorance of the DASH diet rather than any other reason.

#### Content Analyses

Content analyses were performed on the dining menus of twelve nursing home facilities to explore how DASH diet elements were incorporated into the foods of senior residents. This content analysis provided both a quantitative and qualitative look at the prevalence of the DASH diet in nursing homes. The content analyses were separated into two iterations. Iteration one focused on separating menu items that adhered to the DASH diet guidelines, and those that did not. Iteration two focused on tabulating the frequency of each food found in both categories (DASH diet adherent and not).

In iteration one, the coding of the menus was completed by the intramethod approach to triangulation via Renz et al. (2018), the Primary Care Program Director at the Perelman School of Medicine. The food groups are organized by those that align with the DASH diet guidelines (Theme 1) and those that do not (Theme 2). For instance, mixed greens from a winter salad would fall into Theme 1, while deli ham would fall into Theme 2.

In iteration two, the foods found in each theme were tabulated to provide more detailed insight into how each facility incorporates different foods into the same food groups. For example, Facility A provides grilled salmon topped with a garlic herb butter and grilled lemon for non-red and unprocessed protein, while Facility C provides braised chicken with mushroom barley risotto and spinach casserole for non-red and unprocessed protein.

#### Frequency of Each Food Category on Menus, by Facility

In Table 6, each menu incorporated at least one serving of fruits and vegetables. These two food groups were the only groups that were included in every facility’s menu. Whole grains were included in all facility’s menus except for Facility L. Roughly half of the menus included nuts and legumes, as well as low-fat dairy. Three menus included sweetened beverages, which was the food group that was included the least across all menus. Facility G incorporated the highest amount of DASH diet-adherent foods into their menu, while Facility F incorporated the least amount of DASH diet-adherent foods into their menu.

**Table 6.** Based on analysis of nursing home menus.

| Frequency of Food Categories on Facility's Menu | Fruit | Vegetable | Nuts and Legumes | Whole Grains | Low-Fat Dairy | Red and Processed Meats | Sweetened Beverages |
| --- | --- | --- | --- | --- | --- | --- | --- |
| Facility A | 9 | 17 | 2 | 1 | 5 | 6 | 0 |
| Facility B | 5 | 4 | 0 | 1 | 0 | 2 | 0 |
| Facility C | 11 | 26 | 6 | 2 | 0 | 0 | 0 |
| Facility D | 19 | 14 | 0 | 5 | 1 | 18 | 1 |
| Facility E | 11 | 23 | 2 | 3 | 6 | 5 | 6 |
| Facility F | 2 | 1 | 1 | 1 | 0 | 4 | 0 |
| Facility G | 9 | 22 | 1 | 7 | 1 | 2 | 0 |
| Facility H | 4 | 37 | 2 | 1 | 0 | 7 | 0 |
| Facility I | 15 | 21 | 0 | 3 | 7 | 7 | 0 |
| Facility J | 8 | 4 | 0 | 2 | 0 | 2 | 0 |
| Facility K | 6 | 15 | 0 | 1 | 3 | 5 | 4 |
| Facility L | 12 | 28 | 2 | 0 | 7 | 10 | 0 |

#### Frequency and Percentage of Red and Processed Meats on Menu out of Total Meats, Grouped by Facility

**Graph 1.**
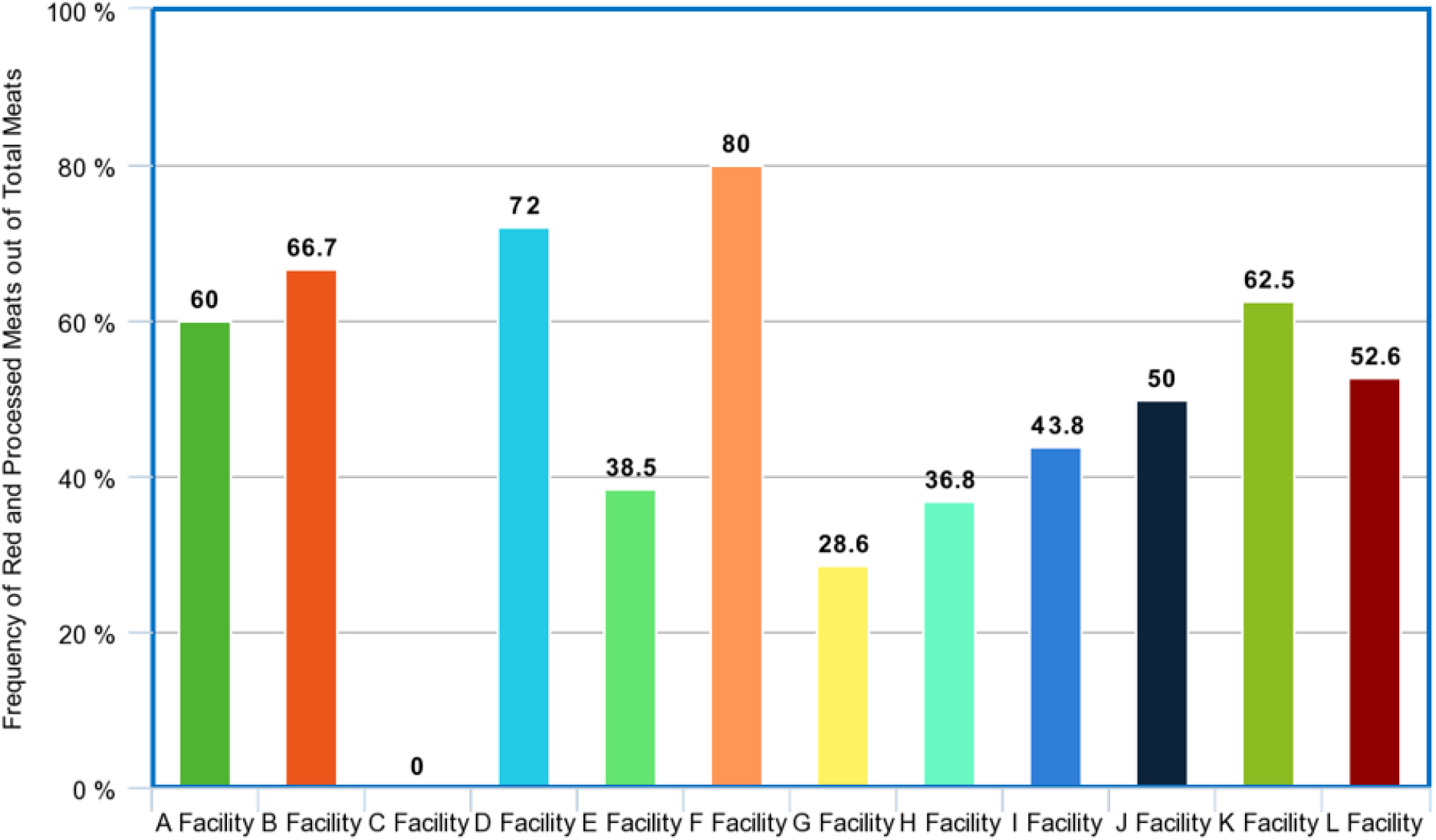
Based on raw data from Table 6.

Red and processed meats was the most prevalent food item across facility menus that did not adhere to the DASH diet. Facility F (80%) had the highest percentage of red and processed meats on their menus compared to total meats (red and processed meats and white and unprocessed meats), while Facility C had the lowest percentage (0%). Facility G (28.6%) had the lowest non-zero percentage of red and processed meats on their menus.

## 6. DISCUSSION

This research study aimed to explore the prevalence of DASH diet elements in Central Texas nursing homes with hypertensive residents.

### Findings

After reviewing the results from both the quantitative and qualitative methods, it can be reasoned that 100% of all the facilities that participated in this study incorporated elements of the DASH diet into their menus and meal plans. Based on table 4, all facilities met at least 30% of the total criteria. In facilities A, B, C, D, and E, this adherence is unintentional, as they responded “no” when asked if they intentionally incorporated the DASH diet into their facility’s menus and meal plans. The intention is unknown for the rest of the nursing homes. This is a limitation, as the other nursing homes did not agree to the interview question, so whether they intentionally incorporate the DASH diet into their meal plans is now known.

Based on the data from table 5, the percentage of facilities (60%) that were highly adherent to the DASH diet was greater than the percentage of facilities (40%) that had low adherence to the DASH diet. When triangulating the data, the data that depicted the extent to which nursing homes incorporate the DASH diet into the menus depended on the method used to investigate this question. When the questionnaire was used, facilities A, C, and E had the highest adherence to the DASH diet, and therefore had the most servings of food that fit into elements of the DASH diet. When performing a content analysis of the menu, these three facilities had the seventh, fourth, and fourth (tied, out of twelve) most menu items that adhered to the DASH diet, respectively. When the questionnaire was used, facilities B and D had the lowest adherence to the DASH diet, and therefore had the lowest number of servings of food that fit into the DASH diet guidelines. When the data analysis was performed, these two facilities had the tenth and sixth most adherence to the DASH diet guidelines (out of twelve). This discrepancy could be attributed to the nutritionist’s faulty reporting of their facility’s food groups, or a change in the seasonal menu that was not mentioned when sent. This finding demonstrates the importance of triangulating the methods. If the study scrutinized data via only one method (questionnaire, interview, content analyses) the data from each facility would either be over or underreported, and therefore not present an accurate view of the nursing home’s meal plans. With the triangulation, both the serving size of each food group and the amount of food that adheres to the DASH diet based on the menu is known. Since all five facilities that responded to the questionnaire stated that they did not intentionally incorporate DASH diet elements into their meal plans, nor were even aware of the DASH diet, there is no correlation to be reported between intentions and data.

A content analysis of the menus concluded that each facility incorporated foods that aligned with the DASH diet guidelines into their meal plans. Although not all foods aligned with the DASH diet guidelines, residents always had an option to choose a meal that aligned with the DASH diet guidelines.

Another important finding was that out of the foods that did not adhere to the DASH diet in facilities, red and processed meats were the most common. Half of the participating facilities had more red and processed meats on their menu than other protein. Claudia L. Campos, an internist at the Atrium Wake Forest Baptist Hospital, argues that since red and processed meats can increase risk of type 2 diabetes, coronary heart disease, and stroke, cutting down on this food group will be beneficial to residents, especially those with hypertension [4].

Another finding that is worth reporting is that the nutritionists at the nursing homes incorporated elements of the DASH diet into the menus without knowing that they aligned with the DASH diet, nor being aware of the benefits of the DASH diet and hypertension. According to the data from the interview, none of the five facilities knew about the DASH diet previous to the questionnaire and none of them reported intentionally including elements of the DASH diet into their meal plans. It is extremely beneficial that nutritionists are incorporating foods that align with the DASH diet into their meal plans for hypertensive residents, but it is also important that they research and acknowledge the health benefits, as including a higher proportion of foods that align with the DASH diet is beneficial to lowering blood pressure, especially in those with high blood pressure.

### Fulfillment of Gaps in the Research

This study addresses multiple gaps in the research. Firstly, previous studies only included hypertensive patients as study subjects. In this study, both hypertensive patients and nutritionists/meal planners were used as subjects. These subjects are important because they overlook and are the most knowledgeable about the culinary and nutritional aspects of the facility, making them the most appropriate subjects for a study about the DASH diet. Secondly, in previous studies, subjects were gathered from a randomized sample of the hypertensive population. This means that these subjects could be teachers, doctors, seniors, etc. My study specifically aims to study the prevalence of the DASH diet in seniors living in nursing home facilities. This decreases the scope and allows for more specific results, as a smaller population within an enclosed facility was studied. Lasty, and most importantly, previous research did not address the extent to which nursing home facilities incorporated elements of the DASH diet into their facility’s meal plans. This was the basis of my research, which filled in other smaller gaps as well, including exploring the most common food group that did not adhere to the DASH diet in facilities.

### Implications

The results of this study can encourage nursing home facilities to begin intentionally incorporating elements of the DASH diet into their facilities. Although most already do, not all have reached a high adherence. If all nursing homes are willing to incorporate enough DASH diet foods to reach high adherence based on the Fung et al. DASH diet scoring method, hypertensive individuals will be benefited in the process. Now that the facilities are aware of what the DASH diet is and how it benefits the health of hypertensive residents, they can provide foods that are more relevant to residents. This study can also prompt nutritionists and meal planners to allocate more of the dining budget to DASH diet adherent ingredients. Doing this will increase the amount of DASH diet adherent food in the menus. Overall, this research can educate nutritionists and meal planners in nursing home facilities, and all hypertensive patients and those taking care of hypertensive patients, about how the DASH diet can reduce blood pressure and promote heart health.

### Limitations

All five facilities that responded to the questionnaire over the phone were extremely busy after the holiday break and were not able to respond to more than one interview question. Had this project occurred at a less hectic time, there would have been more insight into the process nutritionists and meal planners take to plan the residents’ menus and meals. Also, a limitation with the menus is that some facilities provided menus with general food names (wild rice pilaf) and some facilities provided menus with specific ingredients present in the dish (traditional chili with beans, topped with cheddar cheese and onion, served with sour cream and a corn muffin). This made it difficult to track the DASH diet adherence level for specific foods that did not contain specific ingredients. I negated this limitation by analyzing the ingredients present in the title of the dishes that do not include an ingredient list. Also, some menus included breakfast, lunch, and dinner meal options, while some provided only one of the three. Because the latter provided only one meal of the day, it could not capture the full extent of the facility’s meal plans.

### Future Research Areas

The subjects can be expanded to different states beyond Texas, and even globally. This is important because the prevalence of specific diets vary according to the region of the world. Each region of the world has their own culinary taste and style, so different diets will either be less or more common in each region. Other diets that are beneficial to heart health can also be studied. For example, the prevalence of the Mediterranean diet or the Ornish diet can be studied in other facilities around the world.

## Data Availability

All data produced in the present study are available upon reasonable request to the authors.

